# Peri-Procedural Healthcare Costs of Thromboembolic and Bleeding Events in Patients on Direct Oral Anticoagulants Undergoing High-Risk Endoscopy

**DOI:** 10.64898/2026.07.31.26359377

**Authors:** Zachary L. Smith, Nauzer Forbes, B. Joseph Elmunzer, Denise M. Scholtens, Christian T Ruff

**Author notes:** **Corresponding Author** Zachary L. Smith, DO, MSc. 8701 Watertown Plank Rd Hub for Collaborative Medicine, 6th Fl Milwaukee, WI, USA, 53226.

## Abstract

**Background and study aims:** Patients on direct oral anticoagulants (DOACs) undergoing high-risk endoscopy face competing risks of bleeding and thromboembolism during peri-procedural anticoagulant interruption. We quantified peri-procedural adverse event (AE) rates and incremental healthcare costs.

**Patients and methods:** Retrospective cohort study using the TriNetX Linked Claims database (2015–2025), a US-wide multi-payer claims network. Adults on DOAC or warfarin therapy who underwent high-risk endoscopy were included (N=2,486 unique patients; N=1,933 with linked cost data). Peri-procedural bleeding and thromboembolic events were identified using principal-diagnosis codes in acute care encounters (emergency department or inpatient) within 90 days of the index procedure. Unadjusted and multivariable-adjusted incremental 90-day costs were compared between patients with and without each AE type using generalized linear models.

**Results:** Among 2,486 anticoagulated patients (median age 65; 61.5% male; 81% with atrial fibrillation), bleeding occurred in 18.0% at 7 days and 22.9% at 90 days. Stroke/TIA occurred in 0.5% at 7 days and 1.7% at 90 days. Each thromboembolic event cost approximately 2.5 times more than each bleeding event. Stroke/TIA added $8,874 unadjusted (p<0.001) and $5,798 adjusted (cost ratio 1.56; 95% CI 1.11-2.20; p=0.011); bleeding added $3,471 unadjusted and $3,157 adjusted (cost ratio 1.33; 95% CI 1.18-1.50; p<0.001). Patients experiencing both bleeding and subsequent stroke/TIA had the highest costs (mean $23,716).

**Conclusions:** Peri-procedural thromboembolic events cost the healthcare system 2.5-fold more per event than bleeding, underscoring the clinical and economic importance of periprocedural DOAC management and motivating randomized evidence on optimal DOAC resumption timing strategies.

## INTRODUCTION

The use of direct oral anticoagulants (DOACs) has risen substantially since their introduction in 2010^1–3^. Current estimates suggest that nearly 10% of the US population over age 50 receives oral anticoagulation, predominantly for atrial fibrillation (AF)^4^. In parallel, the utilization of interventional endoscopic procedures has expanded as technique innovations have opened pathways to organ-sparing treatments for conditions previously managed surgically^5, 6^. Many of these procedures carry elevated bleeding risk, often exceeding 10%^7–10^.

Current guidelines from the American College of Gastroenterology (ACG) and Canadian Association of Gastroenterology (CAG) acknowledge significant evidence gaps in periprocedural DOAC management, particularly regarding the optimal timing of resumption after high-risk procedures^11^. Most guidelines defer this decision to the treating physician, creating substantial practice variation^12–15^. The clinical consequences of suboptimal DOAC management are bidirectional. Early resumption may increase delayed post-procedural bleeding, while delayed resumption leaves patients at greater risk for thromboembolic events – particularly stroke. Procedure-specific delayed bleeding rates in DOAC patients undergoing high-risk endoscopy are substantial, ranging from approximately 5% for POEM in antithrombotic users to 14–17% for ERCP with sphincterotomy and variceal therapy and 8–23% for endoscopic submucosal dissection^16–20^. Although periprocedural thromboembolic event rates appear low (0.2–0.6%) under short-duration DOAC interruption in landmark studies^7, 21–23^, a post hoc analysis of the ARISTOTLE trial revealed a striking duration-dependent stroke risk – rising from 0.086% with ≤3 days of interruption to 1.0% with ≥8 days^24^ Both categories of adverse events (AEs) carry substantial morbidity, mortality, and healthcare costs. Prior literature has estimated the cost of an ischemic stroke hospitalization at $10,000–$19,000 USD in acute hospital charges alone^25, 26^. However, data characterizing the peri-procedural cost burden of AEs specifically in anticoagulated patients undergoing high-risk endoscopy are lacking.

Understanding these costs is needed to quantify the economic impact of the current evidence gap, support cost-effectiveness evaluation of management strategies, and inform the economic rationale for a proposed large multicenter randomized controlled trial (RCT) comparing different times to resumption of DOACs after high-risk endoscopy. We conducted a national claims-based analysis to quantify adverse event rates and their incremental healthcare costs in patients on DOACs undergoing high-risk endoscopic procedures.

## PATIENTS AND METHODS

### Data Source

We used the TriNetX Linked Claims database, a federated research network containing de-identified electronic health record and linked closed claims data from over 100 million patients in the United States across 44 healthcare organizations. The linked claims component provides pharmacy fills, medical claims with proxy cost data, and enrollment information. The individual patient-level dataset was requested from TriNetX on March 18, 2026. A data use agreement between TriNetX and the Medical College of Wisconsin (MCW) was fully executed on March 25, 2026 and the dataset downloaded in full on March 26, 2026. The MCW Institutional Review Board reviewed this study (PRO00058680) and determined it did not meet criteria for human subjects research at 45 CFR 46.102. This study is reported in accordance with the Strengthening the Reporting of Observational Studies in Epidemiology (STROBE) guideline for cohort studies^27^.

### Study Population

We identified all patients who underwent high-risk endoscopic procedures between 2015 and 2025 using Current Procedural Terminology (CPT) codes (**Supplemental Table 1**). High-risk procedures included endoscopic mucosal resection (EMR), endoscopic retrograde cholangiopancreatography (ERCP) with sphincterotomy, peroral endoscopic myotomy (POEM), endoscopic ultrasound (EUS)-guided transmural drainage, and esophageal variceal band ligation or sclerotherapy. Codes for procedures with snare polypectomy were excluded on the presumption that most codes likely represented cold snare polypectomy (i.e. without diathermy) and therefore, are considered low-risk for delayed bleeding^28, 29^.

We restricted the analysis to patients with an anticoagulant prescription fill (DOAC or warfarin) within 180 days before the index procedure. DOACs were identified by brand-name matching: apixaban (Eliquis), rivaroxaban (Xarelto), dabigatran (Pradaxa), and edoxaban (Savaysa).

Warfarin was identified by brand names Coumadin and Jantoven. Warfarin patients were included to widen the cost-burden lens to all anticoagulated patients undergoing high-risk endoscopy, consistent with the pre-specified protocol AF was identified using ICD-10-CM code I48.x in the patient’s diagnosis history.

### Peri-Procedural Event Definitions

Peri-procedural bleeding and thromboembolic events were identified using ICD-10-CM codes within 90 days of the index procedure. Bleeding events comprised five non-mutually-exclusive subtypes (gastrointestinal, intracranial, genitourinary, post-procedural, and not-elsewhere-classified hemorrhage); patients meeting multiple subtypes were deduplicated in “any bleeding” counts. Thromboembolic events included ischemic stroke, transient ischemic attack, pulmonary embolism, deep vein thrombosis, systemic embolism, and acute myocardial infarction. Complete code lists are provided in **Supplemental Table 2**.

The strict event definition required the diagnosis to appear as the principal diagnosis on an acute-care encounter (emergency department or inpatient), determined by a tiered algorithm of facility indicator, place of service, encounter type, and DRG code presence. Events with a principal diagnosis but unclassifiable setting were retained to avoid excluding facilities with incomplete administrative data. Diagnoses coded on the index-procedure encounter were excluded, except secondary bleeding diagnoses retained to capture post-procedural hemorrhage. The inclusive sensitivity definition is detailed in **Supplemental Table 3**.

### Cost Measurement

Total 90-day peri-procedural costs were computed by summing TriNetX proxy cost values from linked medical claims within 90 days of the index procedure. Proxy cost is a standardized cost estimate derived by TriNetX from CMS Medicare prospective payment system fee schedules, with line-of-business adjustments converting Medicare base rates to commercial and managed Medicaid equivalents (full algorithm available from TriNetX documentation). Proxy costs are designed for relative resource utilization comparisons across patient groups and cannot be interpreted as actual reimbursement amounts. Prescription claims were excluded from the primary cost calculation; reversal agents administered in the hospital setting appear as medical claims and are captured in proxy cost. The primary cost analysis compared 90-day total costs between patients who did and did not experience each AE type.

### Statistical Analysis

Covariate definitions, including CHA2DS2-VASc and HAS-BLED computation from claims-based components, are provided in **Supplemental Table 4**. Baseline characteristics were summarized as medians (IQR) or frequencies (%). The unit of analysis was the patient; each contributed one index event defined by the first qualifying high-risk endoscopic procedure.

Cost comparisons used a generalized linear model with gamma family and log link, appropriate for right-skewed cost data. For each AE type, we fit an unadjusted model and a multivariable-adjusted model including age, sex, CHA2DS2-VASc, HAS-BLED, procedure category, payer type, procedure year, and anticoagulant agent; cost ratios (exp[β]) and incremental costs (marginal effects) with 95% confidence intervals are reported. Pre-specified subgroup analyses examined the AF+DOAC population and stratified cost by procedure type and anticoagulant. An exploratory logistic regression, with and without covariate adjustment, assessed whether peri-procedural bleeding was associated with subsequent stroke or TIA.

Three sensitivity analyses tested robustness: an inclusive event definition accepting any matching ICD-10 diagnosis regardless of encounter setting; restriction to patients with claims data spanning the full 90-day window; and restriction to patients with no enrollment gap >30 days. Baseline characteristics were compared between patients with and without linked cost data to assess potential selection bias (**Supplemental Table 5**). Analyses were performed in R version 4.5.3 (R Foundation for Statistical Computing, Vienna, Austria); a two-sided p<0.05 was considered statistically significant.

## RESULTS

### Study Population

Among 144,790 high-risk endoscopic procedures identified in the database, 2,486 unique patients (2.5%) were receiving anticoagulation therapy (**Supplemental Figure 1**). Apixaban was the most common agent (1,315 patients; 52.9%), followed by rivaroxaban (960; 38.6%), warfarin (165; 6.6%), dabigatran (43; 1.7%), and edoxaban (3; 0.1%). A total of 2,014 patients (81.0%) had documented AF.

The median age was 65 years (IQR 59–73), and 61.5% were male. The median CHA2DS2-VASc score was 4 (IQR 2–5) and median HAS-BLED score was 3 (IQR 2–4). Congestive heart failure was present in 39.9%, hypertension in 85.0%, diabetes in 48.9%, and prior stroke/TIA in 15.0%. Prior bleeding within 1 year was documented in 41.9%, predominantly GI hemorrhage (29.3%) and other hemorrhage (20.4%), with a median of 28 days (IQR 2-107) before the index procedure. Just over half (51.1%) of prior bleeding events occurred within 30 days of the procedure (**Supplemental Table 6**). Of these, the majority were patients undergoing variceal therapy (35.2%). The most common procedures were EMR (44.4%), ERCP with sphincterotomy (32.9%), variceal band ligation (17.0%), and transmural drainage (4.6%). The majority of procedures (69.4%) were performed in the inpatient setting (**Table 1**).

**Table 1.** Baseline Characteristics of Anticoagulated Patients Undergoing High-Risk Endoscopy (N = 2,486)

| Characteristic | Value |
| --- | --- |
| Age, years, median [IQR] | 65 [59, 73] |
| Male sex, n (%) | 1,529 (61.5) |
| **Race/ethnicity, n (%)** |  |
| White | 1,772 (71.3) |
| Black or African American | 341 (13.7) |
| Asian | 96 (3.9) |
| Other/Unknown | 277 (11.1) |
| **Payer type, n (%)** |  |
| Commercial | 945 (38.0) |
| Medicare Advantage | 857 (34.5) |
| Medicaid | 587 (23.6) |
| Unknown | 48 (1.9) |
| **Comorbidities** |  |
| CHA2DS2-VASc score, median [IQR] | 4 [2, 5] |
| HAS-BLED score, median [IQR] | 3 [2, 4] |
| Atrial fibrillation, n (%) | 2,014 (81.0) |
| Congestive heart failure, n (%) | 992 (39.9) |
| Hypertension, n (%) | 2,113 (85.0) |
| Diabetes mellitus, n (%) | 1,215 (48.9) |
| Prior stroke/TIA, n (%) | 373 (15.0) |
| Vascular disease, n (%) | 1,471 (59.2) |
| Chronic kidney disease, n (%) | 1,015 (40.8) |
| Chronic liver disease, n (%) | 1,010 (40.6) |
| Alcohol use disorder, n (%) | 358 (14.4) |
| Prior bleeding (1 year), n (%) | 1,042 (41.9) |
| Antiplatelet use (90 days), n (%) | 37 (1.5) |
| **Anticoagulant agent, n (%)** |  |
| Apixaban | 1,315 (52.9) |
| Rivaroxaban | 960 (38.6) |
| Warfarin | 165 (6.6) |
| Dabigatran | 43 (1.7) |
| Edoxaban | 3 (0.1) |
| **Procedure category, n (%)** |  |
| Endoscopic mucosal resection | 1,105 (44.4) |
| ERCP with sphincterotomy | 818 (32.9) |
| Variceal band ligation/sclerotherapy | 423 (17.0) |
| Transmural drainage | 114 (4.6) |
| Peroral endoscopic myotomy | 26 (1.0) |
| **Procedure setting, n (%)** |  |
| Inpatient | 1,725 (69.4) |
| Outpatient/ASC | 321 (12.9) |
| Emergency department | 42 (1.7) |
| Unknown | 398 (16.0) |

*IQR = interquartile range; ERCP = endoscopic retrograde cholangiopancreatography; ASC = ambulatory surgery center.*

### Peri-Procedural Adverse Event Rates

Using the strict event definition, the 7-day bleeding rate was 18.0% and the 7-day thromboembolic rate was 1.9%. Stroke/TIA specifically occurred in 0.5% at 7 days. At 30 days, bleeding increased to 20.5%, thromboembolic events to 3.7%, and stroke/TIA to 0.8%. At 90 days, 22.9% experienced bleeding, 6.2% experienced a thromboembolic event, and 1.7% had a stroke or TIA (**Table 2, Supplemental Figure 2**).

**Table 2.** Peri-Procedural Event Rates in Anticoagulated Patients by Time Window (N = 2,486)

| Event | 7 Days | 14 Days | 30 Days | 90 Days |
| --- | --- | --- | --- | --- |
| Bleeding | 448 (18.0) | 486 (19.5) | 510 (20.5) | 569 (22.9) |
| Any thromboembolic | 48 (1.9) | 66 (2.7) | 91 (3.7) | 153 (6.2) |
| Stroke/TIA | 13 (0.5) | 15 (0.6) | 21 (0.8) | 43 (1.7) |

*Values are n (%). Event ascertainment used a strict definition requiring principal diagnosis coding in an acute care setting. TIA = transient ischemic attack.*

Among 90-day thromboembolic events, DVT was most common (2.0%), followed by PE (1.5%), ischemic stroke (1.4%), MI (1.2%), TIA (0.4%), and systemic arterial embolism (0.4%) (**Table 3**). GI hemorrhage accounted for the majority of bleeding events (19.6%), with post-procedural hemorrhage in 2.1% and intracranial hemorrhage in 0.4% (**Table 3**). Bleeding event rates by subtype at 7, 30, and 90 days are provided in **Supplemental Table 7**.

**Table 3.** Peri-Procedural Event Subtypes at 90 Days in Anticoagulated Patients (N = 2,486)

| Category | Subtype | Events | Rate (%) |
| --- | --- | --- | --- |
| Bleeding | GI hemorrhage | 487 | 19.6 |
|  | Other hemorrhage | 224 | 9.0 |
|  | Post-procedural | 51 | 2.1 |
|  | GU bleeding | 43 | 1.7 |
|  | Intracranial hemorrhage | 9 | 0.4 |
| Thromboembolic | Deep vein thrombosis | 49 | 2.0 |
|  | Pulmonary embolism | 38 | 1.5 |
|  | Ischemic stroke | 35 | 1.4 |
|  | Myocardial infarction | 30 | 1.2 |
|  | Systemic embolism | 9 | 0.4 |
|  | TIA | 9 | 0.4 |

*GI = gastrointestinal; GU = genitourinary; TIA = transient ischemic attack. Patients may have multiple event subtypes.*

In the AF+DOAC subgroup (n=1,911), the rate of stroke/TIA at 7, 30, and 90 days was 0.5% (10 patients), 0.8% (16 patients), and 1.8% (36 patients), respectively.

### Incremental Cost of Peri-Procedural Adverse Events

Among the 1,933 anticoagulated patients with linked 90-day cost data, the overall mean 90-day peri-procedural cost was $10,407 (median $7,193; IQR $3,823–$12,405) (**Figure 1**). Costs varied by procedure type, with transmural drainage highest (mean $15,147) and POEM lowest ($7,709) (**Supplemental Table 8**).

**Figure 1.**
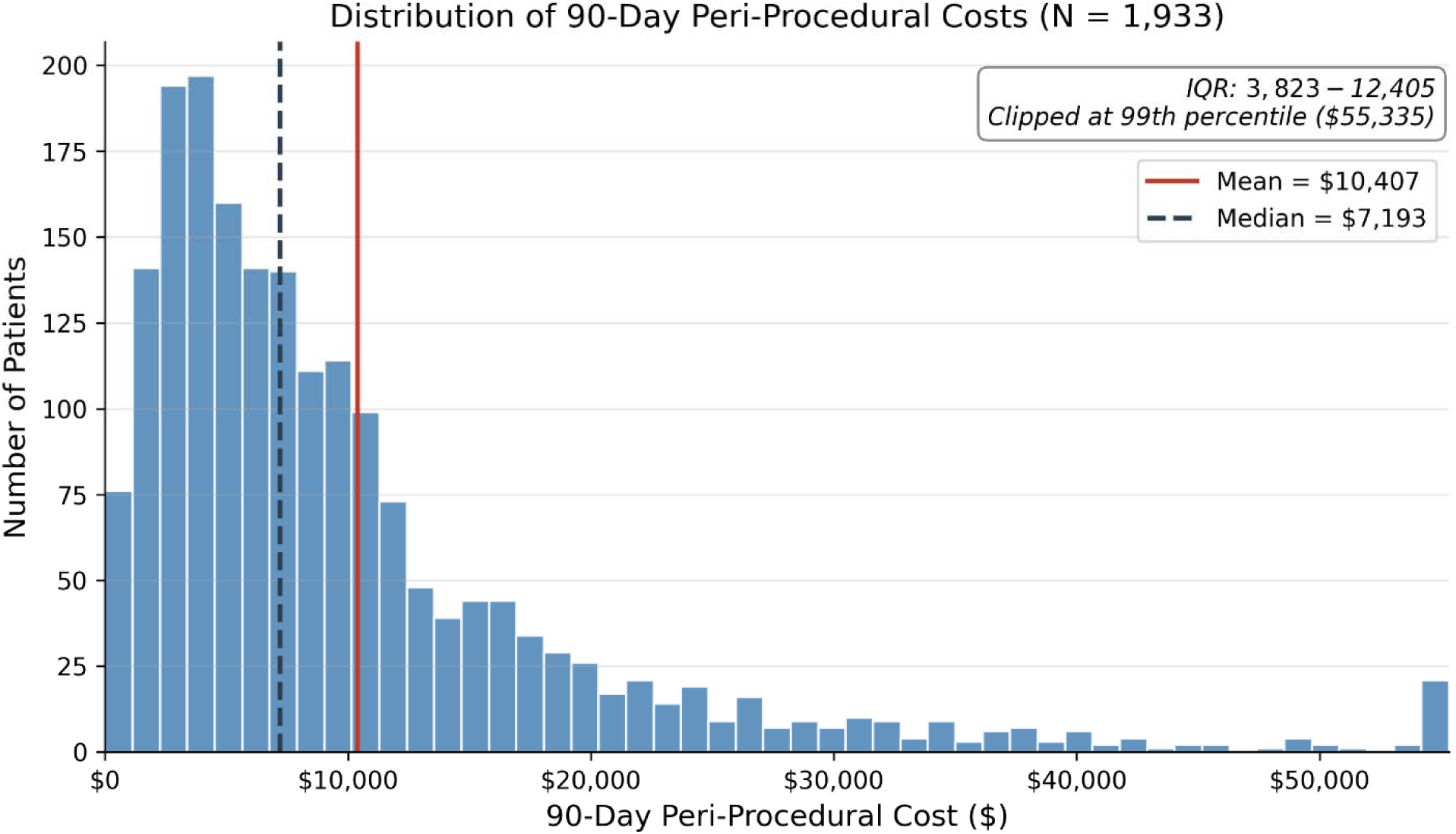
Distribution of 90-day peri-procedural costs in anticoagulated patients undergoing high-risk endoscopy.

Stroke and TIA carried an incremental cost of $8,874 per patient (mean $19,127 with vs. $10,253 without; p<0.001) (**Supplemental Table 9, Figure 2**). Of these 37 patients, 31 had only a stroke code (mean cost $16,663) and 7 had only a TIA code (mean cost $30,321; 1 patient had both). Two TIA cases ($66,504 and $40,406) accounted for 51% of group costs, supporting the grouping of stroke/TIA into one event definition.

**Figure 2.**
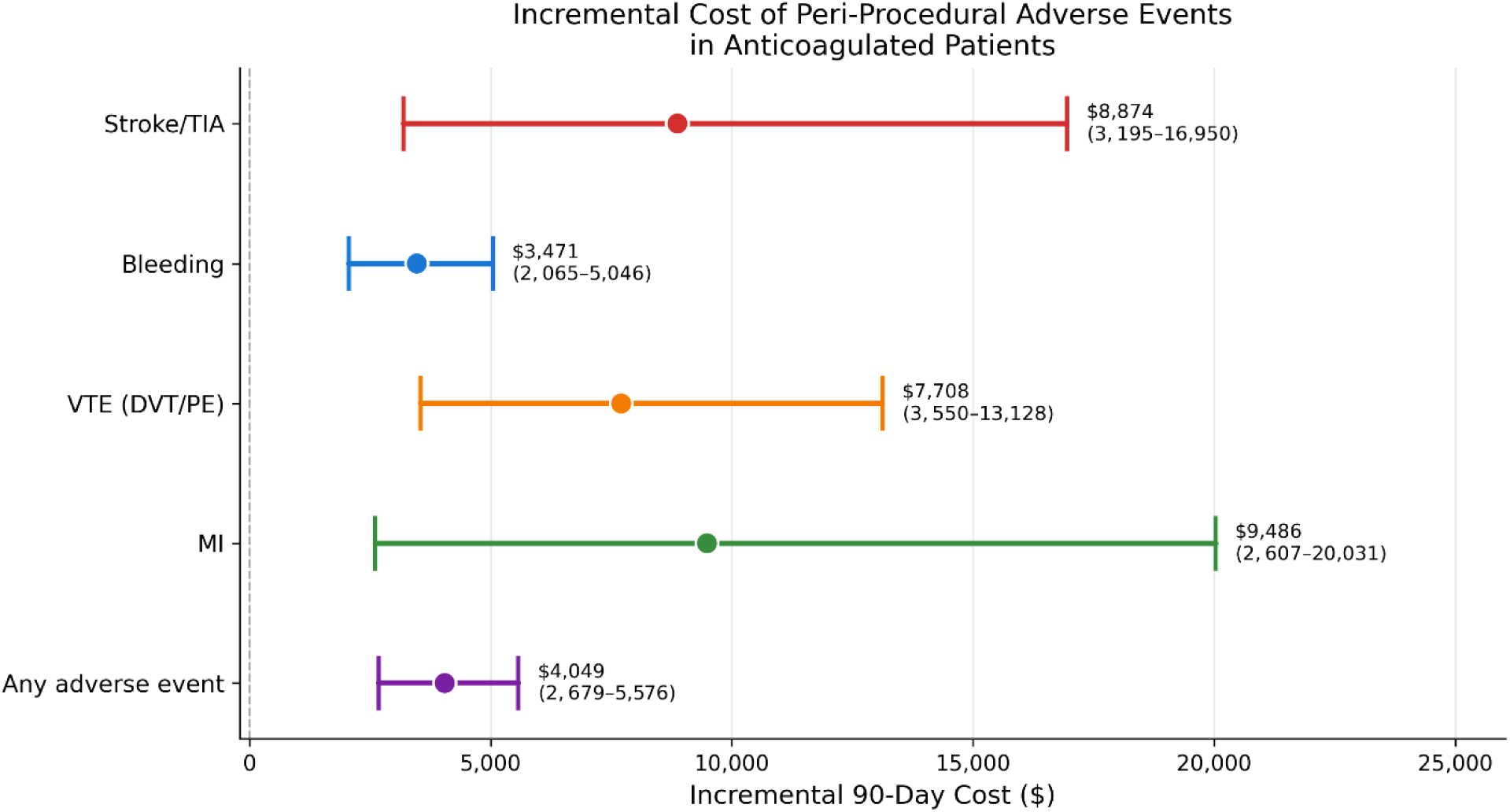
Incremental 90-day cost of peri-procedural adverse events in anticoagulated patients. Points represent the unadjusted incremental mean cost (mean cost with adverse event minus mean cost without); horizontal lines represent 95% confidence intervals derived from a gamma-family generalized linear model with log link.

Other VTE events (e.g. deep venous thrombosis, pulmonary embolism) added $7,708 in incremental costs (mean $17,867 vs. $10,159; p<0.001). Myocardial infarction carried the highest per-event increment at $9,486 (mean $19,786 vs. $10,300; p=0.003), though the small sample (n=25) limits precision (**Figure 2**).

Bleeding, despite being the most common AE (n=457; 23.6%), added the least per event at $3,471 (mean $13,072 vs. $9,601; p<0.001). Each thromboembolic event cost approximately 2.5 times more than each bleeding event. Any AE (n=529; 27.4% of patients; patients may contribute to multiple AE categories) added $4,049 (mean $13,362 vs. $9,313; p<0.001).

In the subgroup of patients with AF on DOAC therapy, stroke and TIA carried an incremental cost of $9,698 (mean $20,114 vs. $10,416). Bleeding added $3,694 and VTE added $8,676 (**Supplemental Table 10**). Results by anticoagulant agent are presented in **Supplemental Table 11**.

### Exploratory Analysis: Bleeding and Subsequent Stroke/TIA Risk

In an exploratory analysis, patients who experienced bleeding within 30 days of their procedure had a numerically higher 90-day stroke and/or TIA rate compared with those who did not bleed (2.7% vs. 1.5%; OR 1.90; 95% CI 0.99–3.61), though this was not statistically significant.

Among the 13 patients who experienced stroke/TIA temporally after a bleeding event, the median interval from bleed to stroke was 6 days (mean 22.4 days). Patients who experienced both bleeding and stroke/TIA had the highest 90-day costs (mean $23,716), representing an incremental $11,651 above bleed-only patients (Supplemental Table 12). However, after multivariable adjustment for age, sex, CHA2DS2-VASc, HAS-BLED, procedure year, and anticoagulant agent, the association between bleeding and subsequent stroke/TIA was attenuated and crossed the null (adjusted OR 1.19; 95% CI 0.60-2.38; p=0.62). CHA2DS2-VASc score was the strongest predictor of stroke/TIA (OR 1.58 per point; 95% CI 1.24-2.02; p<0.001), suggesting that the unadjusted association was driven by confounding from baseline thromboembolic risk (**Supplemental Table 12**).

Results were robust across sensitivity analyses. Restricting to patients with claims spanning the full 90-day window (87.4% of patients with cost data) yielded incremental costs within 10% of the primary estimates (stroke/TIA: $8,034 vs $8,874; bleeding: $3,509 vs $3,471). Only 11 patients had enrollment gaps exceeding 30 days within the 90-day window; restricting to continuously enrolled patients did not meaningfully change results.

## DISCUSSION

This national claims-based study provides a comprehensive characterization of peri-procedural AE rates and their healthcare costs in anticoagulated patients undergoing high-risk endoscopy. The central finding is that each thromboembolic event costs approximately 2.5 times more than each bleeding event ($8,874 vs. $3,471 for stroke/TIA vs. bleeding). This cost asymmetry has direct implications for how clinicians balance bleeding and thromboembolic risk when managing periprocedural anticoagulation.

The clinical rationale for optimizing DOAC resumption timing has been well established. Current guidelines acknowledge the evidence gap^11, 15^, and a recent survey demonstrated substantial practice variation among endoscopists regarding the approach to DOAC resumption after high-risk endoscopic procedures^30, 31^. The present study adds an economic dimension to this discussion. If early DOAC resumption after high-risk endoscopy modestly increases bleeding risk but meaningfully reduces thromboembolic risk, such a strategy could be cost-neutral or even cost-saving on a population level given the 2.5-fold difference between these AEs. A rigorous, well-powered randomized trial comparing various times to DOAC resumption after high-risk endoscopic procedures is the best way to answer this clinically and economically critical question.

The observed 7-day stroke/TIA rate of 0.5% and 90-day rate of 1.7% in our anticoagulated population are consistent with prior estimates from both observational and randomized data. In the ARISTOTLE sub-analysis of AF patients undergoing invasive procedures with DOAC interruption, the 30-day stroke incidence was 0.086% with ≤3 days of interruption, 0.61% with 5–7 days, and 1.0% with ≥8 days^24^. The 30-day stroke/TIA rate of 0.8% in the present study falls between the 5–7 day and ≥8 day ARISTOTLE strata, consistent with the typical real-world practice of holding DOACs for ≥5 days around high-risk endoscopic procedures. A study of AF patients undergoing endoscopy reported periprocedural stroke rates of 1.1% per procedure, with rates as high as 2.9% in complex patients^32^. Our strict event definition, which requires principal diagnosis coding in an acute care setting, was specifically designed to exclude prevalent diagnoses that were carried forward from prior encounters. The validity of this approach is supported by the observation that the inclusive sensitivity analysis yielded thromboembolic rates approximately 4-fold higher, confirming that the majority of excluded events were not new acute occurrences.

The 19.6% 90-day GI bleeding rate (**Supplemental Table 7**) is consistent with the procedure mix in our cohort. The population included patients undergoing variceal band ligation, EMR, and ERCP with sphincterotomy, each of which carries baseline delayed bleeding risks of 5–25^7–10^.

Importantly, the per-event proxy cost of bleeding ($3,471) was substantially lower than that of thromboembolic events, suggesting that most bleeding episodes are managed conservatively through observation or repeat endoscopy rather than requiring surgical intervention or prolonged hospitalization. Patients undergoing variceal therapy had the highest observed bleeding rate (52.3% at 90 days). This likely reflects that many variceal banding procedures were performed to treat active or recent variceal hemorrhage rather than as elective prophylaxis, conflating the procedural indication with the post-procedural event. A sensitivity analysis excluding the 222 patients with documented bleeding only within 0–13 days before the procedure (likely representing variceal-hemorrhage indications) reduced the variceal-subgroup bleeding rate to 44.1% and preserved all primary findings, including the 2.5-fold cost asymmetry between thromboembolic and bleeding events. We retained these patients in the primary cohort because the cost-burden analysis — the central question of this study — does not depend on procedural indication; a subsequent thromboembolic event imposes its cost regardless of why the index procedure was performed.

The population-level cost implications of these findings are substantial. Approximately 4 million patients in the United States currently receive DOAC therapy, and roughly 20% undergo an elective or urgent procedure each year, translating to approximately 800,000 periprocedural DOAC management episodes annually^21^. In the PAUSE study, 33.5% of procedures were classified as high-bleeding-risk^23^, translating to approximately 268,000 high-risk procedures in DOAC patients per year. While the total number of high-risk GI endoscopic procedures in anticoagulated patients is not precisely known, even conservative estimates suggest tens of thousands of such encounters per year. At the incremental costs observed in our study, the aggregate national cost burden of peri-procedural AEs in this population likely reaches tens of millions of dollars annually in excess acute healthcare expenditures alone. Globally, approximately 6 million patients with AF require periprocedural anticoagulant management each year, suggesting that the international cost burden is proportionally larger^23^.

Several limitations should be acknowledged. Most importantly, claims data lack the granularity to identify the exact duration of anticoagulant interruption around each procedure. This is a critical limitation because the thromboembolic risk observed in this study inherently stems from the period of anticoagulant interruption, and the duration of that interruption is the modifiable factor that a well-powered randomized trial should be designed to evaluate. Our findings quantify the cost consequences of the current state of practice but cannot attribute them to specific interruption durations. Second, proxy cost data represent standardized estimates rather than actual reimbursement amounts, though relative comparisons remain valid. Third, 22.2% of patients lacked linked cost data, which may introduce selection bias toward patients with claims-based rather than EHR-only records. Fourth, procedure setting could not be determined for 16.0% of patients despite our tiered classification algorithm. An outpatient/elective subgroup analysis (n=504) demonstrated lower AE rates than the overall cohort (**Supplemental Table 13**). Fifth, the 90-day cost window captures the acute burden but not long-term stroke-related disability, rehabilitation, or lost productivity, meaning our estimates may underestimate the true economic impact of peri-procedural stroke. Sixth, concomitant antiplatelet use was identified in only 1.5% of anticoagulated patients and the true antiplatelet exposure is likely underestimated because aspirin, the most commonly used antiplatelet agent, is predominantly purchased over the counter. While 9.8% of our cohort had an antiplatelet fill documented at some point in their claims history, only those with fills within 90 days of the index procedure were captured. Finally, the exploratory association of thromboembolic events after bleeding in this study is underpowered and subject to confounding by severity of illness.

Despite these limitations, this study has several notable strengths. To our knowledge, this is the first study to directly measure and compare the incremental healthcare costs of peri-procedural bleeding and thromboembolic events in anticoagulated patients undergoing high-risk endoscopy. The use of linked claims data, rather than charges or cost-to-charge ratios from a single institution, provides a nationally representative cost perspective. Additionally, TriNetX data have been shown to be broadly generalizable to the US population with respect to age, sex, race, and geographic distribution, particularly among the healthcare-seeking population^33^. Our strict event definition distinguishes new acute events from prevalent diagnoses carried forward from prior encounters, addressing a well-recognized source of outcome misclassification in claims-based studies. The analysis captures costs across all payer types, avoiding the single-payer bias inherent in Medicare-only or Medicaid-only datasets. Finally, the exploratory bleeding-thrombosis analysis, while underpowered and attenuated on adjusted analysis, generates a testable hypothesis that is now supported by the prospective INTERBLEED data^34^ and can be evaluable in a future randomized trial.

In conclusion, peri-procedural AEs in anticoagulated patients undergoing high-risk endoscopy carry a substantial and quantifiable cost burden, with each thromboembolic event costing the healthcare system approximately 2.5 times more than each bleeding event. Extrapolated nationally, the aggregate excess cost of these AEs likely reaches tens of millions of dollars annually, a figure that does not account for long-term disability, rehabilitation, or lost productivity. Considering the global burden of AF and DOAC utilization – the economic scale of this clinical scenario is exceptional. These data underscore that periprocedural DOAC management is not only a clinical problem but also a major economic one. The optimal resumption strategy must rigorously balance the competing risks of bleeding and thromboembolism. The RESUME trial, a large pragmatic randomized controlled trial comparing early versus delayed DOAC resumption after high-risk endoscopy, is designed to provide this evidence and has the potential to reduce both the clinical and economic burden of peri-procedural AEs for the millions of patients globally who require anticoagulation management around endoscopic procedures each year.

## Article Information

### Author Contributions

Zachary L. Smith: Conceptualization, Data curation, Formal analysis, Funding acquisition, Investigation, Methodology, Project administration, Software, Validation, Visualization, Writing – original draft, Writing – review & editing. Nauzer Forbes: Conceptualization, Writing – review & editing. B. Joseph Elmunzer: Conceptualization, Writing – review & editing. Denise M. Scholtens: Methodology, Validation, Writing – review & editing. Christian T Ruff: Conceptualization, Methodology, Writing – review & editing.

### Data Sharing Statement

The study was pre-registered on Open Science Framework (https://doi.org/10.17605/OSF.IO/RK28Q). The analytic code is publicly available at the same registry. Due to the terms of the data use agreement between TriNetX and Medical College of Wisconsin, the individual patient-level dataset cannot be shared publicly.

