## Supplemental Material for "Peri-Procedural Healthcare Costs of Thromboembolic and Bleeding Events in Patients on Direct Oral Anticoagulants Undergoing High-Risk Endoscopy"

**Table of Contents**

Supplemental Table 1. CPT Codes for High-Risk Endoscopic Procedure Definitions

Supplemental Table 2. ICD-10-CM Codes for Bleeding and Thromboembolic Event Definitions

Supplemental Table 3. Sensitivity Analysis: Peri-Procedural Event Rates Using Inclusive Definition vs. Strict Definition (N = 2,486)

Supplemental Table 4. CHA2DS2-VASc and HAS-BLED Score Computation from Claims Data

Supplemental Table 5. Characterization of Missing Cost Data

Supplemental Table 6. Characterization of Prior Bleeding Events in the 365-Day Lookback Period (N = 2,486)

Supplemental Table 7. Bleeding Events by Subtype and Time Window in Anticoagulated Patients (N = 2,486)

Supplemental Table 8. 90-Day Peri-Procedural Cost by Procedure Category in Anticoagulated Patients (Patient Level)

Supplemental Table 9. Unadjusted and Adjusted Incremental 90-Day Cost of Peri-Procedural Adverse Events

Supplemental Table 10. Incremental 90-Day Cost of Adverse Events in the AF+DOAC Subgroup (N = 1,545 with Cost Data)

Supplemental Table 11. 90-Day Peri-Procedural Cost by Anticoagulant Agent

Supplemental Table 12. Exploratory Analysis: Association Between Peri-Procedural Bleeding and Subsequent Stroke/TIA

Supplemental Table 13. Outpatient/Elective Subgroup: Peri-Procedural Event Rates and Costs (Anticoagulated Patients, N = 504)

Supplemental Table 14. Multivariable GLM Coefficients for 90-Day Total Cost

Supplemental Figure 1. Patient flow diagram

Supplemental Figure 2. Peri-procedural event rates in anticoagulated patients by time window (7, 30, and 90 days) using the strict event definition.

**Supplemental Table 1.** CPT Codes for High-Risk Endoscopic Procedure Definitions

| Procedure Category | CPT Code | Description |
|---|---|---|
| EMR | 43254 | EGD with endoscopic mucosal resection |
| | 45390 | Colonoscopy with endoscopic mucosal resection |
| | 45349 | Sigmoidoscopy with endoscopic mucosal resection |
| | 43211 | Esophagoscopy with endoscopic mucosal resection |
| ERCP | 43262 | ERCP with sphincterotomy |
| POEM | 43497 | Peroral endoscopic myotomy |
| Drainage | 43240 | EGD with transmural pseudocyst drainage |
| Variceal | 43244 | EGD with variceal band ligation |
| | 43243 | EGD with variceal injection sclerosis |
| | 43285 | Esophagoscopy with variceal band ligation |
| | 43284 | Esophagoscopy with variceal injection sclerosis |

*CPT = Current Procedural Terminology; EGD = esophagogastroduodenoscopy; EMR = endoscopic mucosal resection; ERCP = endoscopic retrograde cholangiopancreatography; POEM = peroral endoscopic myotomy.*

**Supplemental Table 2.** ICD-10-CM Codes for Bleeding and Thromboembolic Event Definitions

### Bleeding Events

| Category | ICD-10-CM Code(s) | Description |
|---|---|---|
| GI hemorrhage | K92.0, K92.1, K92.2 | Hematemesis, melena, GI hemorrhage unspecified |
| | K62.5 | Hemorrhage of anus and rectum |
| | K25.0, K25.2, K25.4, K25.6 | Gastric ulcer with hemorrhage |
| | K26.0, K26.2, K26.4, K26.6 | Duodenal ulcer with hemorrhage |
| | K27.0, K27.2, K27.4, K27.6 | Peptic ulcer with hemorrhage |
| | K55.21 | Angiodysplasia with hemorrhage |
| | K57.11, K57.13, K57.31, K57.33 | Diverticular disease with hemorrhage |
| | K57.51, K57.53, K57.91, K57.93 | Diverticular disease with hemorrhage (cont.) |
| Intracranial | I60.x, I61.x, I62.x | Subarachnoid, intracerebral, other intracranial hemorrhage |
| Genitourinary | N93.x | Abnormal uterine bleeding |
| | R31.x | Hematuria |
| Post-procedural | K91.84, K91.840, K91.841 | Post-procedural hemorrhage of GI system |
| Other | R58 | Hemorrhage not elsewhere classified |
| | D62 | Acute posthemorrhagic anemia |

### Thromboembolic Events

| Category | ICD-10-CM Code(s) | Description |
|---|---|---|
| Ischemic stroke | I63.x | Cerebral infarction |
| TIA | G45.x | Transient cerebral ischemic attacks |
| Systemic embolism | I74.x | Arterial embolism and thrombosis |
| MI | I21.x | Acute myocardial infarction |
| PE | I26.x | Pulmonary embolism |
| DVT | I82.4x | DVT of deep veins of lower extremity |

*All codes used prefix matching (e.g., I63.x captures I63.0 through I63.9 and all sub-codes). GI = gastrointestinal; TIA = transient ischemic attack; MI = myocardial infarction; PE = pulmonary embolism; DVT = deep vein thrombosis.*

**Supplemental Table 3.** Sensitivity Analysis: Peri-Procedural Event Rates Using Inclusive Definition vs. Strict Definition (N = 2,486)

| Event | Strict 7d | Inclusive 7d | Strict 30d | Inclusive 30d | Strict 90d | Inclusive 90d |
|---|---|---|---|---|---|---|
| Bleeding | 448 (18.0) | 489 (19.7) | 510 (20.5) | 600 (24.1) | 569 (22.9) | 733 (29.5) |
| Any TE | 48 (1.9) | 221 (8.9) | 91 (3.7) | 373 (15.0) | 153 (6.2) | 550 (22.1) |
| Stroke/TIA | 13 (0.5) | 59 (2.4) | 21 (0.8) | 103 (4.1) | 43 (1.7) | 179 (7.2) |

*Values are n (%). Strict definition requires principal diagnosis coding in an acute care setting. Inclusive definition accepts any matching ICD-10 diagnosis in the peri-procedural window regardless of principal diagnosis indicator or encounter setting. The 4-fold difference in thromboembolic event rates between definitions confirms that the majority of events excluded by the strict definition were prevalent diagnoses carried forward on routine encounters rather than new acute events. TE = thromboembolic; TIA = transient ischemic attack.*

**Supplemental Table 4.** CHA2DS2-VASc and HAS-BLED Score Computation from Claims Data

**Panel A:** CHA2DS2-VASc Score

| Component | ICD-10-CM Codes | Points |
|---|---|---|
| Congestive heart failure | I50.x | +1 |
| Hypertension | I10, I11, I12, I13, I14, I15, I16 | +1 |
| Age 65-74 years | From year_of_birth in patient table | +1 |
| Age >= 75 years | From year_of_birth in patient table | +2 |
| Diabetes mellitus | E08, E09, E10, E11, E12, E13 | +1 |
| Prior stroke/TIA/SE | I63, G45, I74 | +2 |
| Vascular disease | I21-I25, I70-I79 | +1 |
| Female sex | Sex field in patient table | +1 |

**Panel B:** Modified HAS-BLED Score

| Component | ICD-10-CM Codes / Source | Points |
|---|---|---|
| Hypertension | I10-I16 | +1 |
| Renal disease | N17, N18, N19 | +1 |
| Liver disease | K70-K77 | +1 |
| Prior stroke | I63, G45, I74 | +1 |
| Prior bleeding | All bleeding ICD-10 codes (Supplemental Table 2) | +1 |
| Age > 65 years | From year_of_birth | +1 |
| Alcohol use disorder | F10.x | +1 |
| Antiplatelet use | Brand matching in medication_drug (90-day lookback) | +1 |
| Labile INR | Not measurable in claims data | Omitted |

*All comorbidities ascertained from diagnosis codes in the 365 days preceding the index procedure. Age calculated as procedure year minus year_of_birth. Maximum possible CHA2DS2-VASc score = 9. Maximum possible modified HAS-BLED score = 8 (labile INR omitted).*

**Supplemental Table 5.** Characterization of Missing Cost Data

| Variable | With Cost (n=1,933) | Without Cost (n=553) | P value |
|---|---|---|---|
| Age, mean years | 63.9 | 68.1 | <0.001 |
| Male, % | 61.5 | 61.7 | 0.41 |
| CHA2DS2-VASc, mean | 3.8 | 3.7 | 0.31 |
| Commercial payer, % | 35.6 | 46.5 | <0.001 |
| EMR, % | 44.4 | 44.7 | 0.91 |
| ERCP, % | 31.9 | 36.5 | 0.04 |
| Variceal therapy, % | 18.3 | 12.5 | 0.001 |

*P values from Mann-Whitney U test (continuous) or chi-square test (categorical). Patients without linked cost data were older and more likely to have commercial insurance. CHA2DS2-VASc and sex were similar between groups.*

**Supplemental Table 6.** Characterization of Prior Bleeding Events in the 365-Day Lookback Period (N = 2,486)

**Panel A:** Prior Bleeding by Type

| Bleeding Type | Patients | Rate (%) |
|---|---|---|
| GI hemorrhage | 728 | 29.3 |
| Other hemorrhage (R58, D62) | 506 | 20.4 |
| Genitourinary bleeding | 260 | 10.5 |
| Post-procedural hemorrhage | 32 | 1.3 |
| Intracranial hemorrhage | 39 | 1.6 |
| **Any prior bleeding** | **1,042** | **41.9** |

**Panel B:** Timing of Most Recent Prior Bleeding Relative to Index Procedure

| Metric | Value |
|---|---|
| Index events with prior bleeding | 1,042 |
| Mean days before procedure | 70 |
| Median days before procedure (IQR) | 28 (2-107) |
| Within 30 days of procedure | 532 (51.1%) |
| Within 90 days | 738 (70.8%) |
| Within 180 days | 900 (86.4%) |

*Prior bleeding defined as any ICD-10-CM bleeding diagnosis code in the 365 days before the index procedure. Patients may have multiple bleeding types. Unit of analysis is the patient (N=2,486). GI = gastrointestinal.*

**Supplemental Table 7.** Bleeding Events by Subtype and Time Window in Anticoagulated Patients (N = 2,486)

| Bleeding Subtype | 7 Days | 30 Days | 90 Days |
|---|---|---|---|
| Gastrointestinal hemorrhage | 402 (16.2) | 453 (18.2) | 487 (19.6) |
| Other hemorrhage | 182 (7.3) | 208 (8.4) | 224 (9.0) |
| Post-procedural hemorrhage | 31 (1.2) | 48 (1.9) | 51 (2.1) |
| Genitourinary bleeding | 17 (0.7) | 22 (0.9) | 43 (1.7) |
| Intracranial hemorrhage | 2 (0.1) | 5 (0.2) | 9 (0.4) |
| **Any bleeding (total)** | **448 (18.0)** | **510 (20.5)** | **569 (22.9)** |

*Values are n (%). Event ascertainment used a strict definition requiring principal diagnosis coding in an acute care setting. Patients may contribute to multiple subtypes. Post-procedural hemorrhage (K91.84x) and gastrointestinal hemorrhage (K92.x and lesion-specific codes) are non-mutually-exclusive; ~76% of patients meeting the PPH criterion also carry a K92.x code, typically on the same encounter.*

**Supplemental Table 8.** 90-Day Peri-Procedural Cost by Procedure Category in Anticoagulated Patients (Patient Level)

| Procedure | N | Mean Cost | Median Cost | IQR | Bleeding Rate (%) |
|---|---|---|---|---|---|
| Transmural drainage | 84 | $15,147 | $10,282 | $6,025–$20,018 | 22.6% |
| ERCP w/ sphincterotomy | 616 | $11,872 | $8,632 | $4,370–$14,909 | 15.7% |
| Variceal therapy | 354 | $10,787 | $7,306 | $3,748–$14,391 | 52.3% |
| EMR | 858 | $8,801 | $6,246 | $3,556–$9,956 | 17.9% |
| POEM | 21 | $7,709 | $3,257 | $1,781–$11,050 | 9.5% |

*All costs in US dollars (proxy cost). IQR = interquartile range; ERCP = endoscopic retrograde cholangiopancreatography; EMR = endoscopic mucosal resection; POEM = peroral endoscopic myotomy.*

### Supplemental Table 9. Unadjusted and Adjusted Incremental 90-Day Cost of Peri-Procedural Adverse Events

| Adverse Event | N | Unadj. CR (95% CI) | Unadj. Incr. | Adj. CR (95% CI) | Adj. Incr. |
|---|---|---|---|---|---|
| Stroke/TIA | 37 | 1.87 (1.31–2.65) | +$8,874 | 1.56 (1.11–2.20) | +$5,798 |
| VTE (DVT/PE) | 66 | 1.76 (1.35–2.29) | +$7,708 | 1.67 (1.29–2.16) | +$6,833 |
| MI | 25 | 1.92 (1.25–2.94) | +$9,486 | 2.16 (1.43–3.26) | +$11,979 |
| Bleeding | 457 | 1.36 (1.22–1.53) | +$3,471 | 1.33 (1.18–1.50) | +$3,157 |
| Any | 529 | 1.43 (1.29–1.60) | +$4,049 | 1.40 (1.25–1.56) | +$3,727 |

*Unadj. CR = unadjusted cost ratio from gamma-family GLM with log link (event indicator only). Adj. CR = adjusted cost ratio from multivariable GLM (gamma, log link) with covariates: age, sex, CHA2DS2-VASc, HAS-BLED, procedure category, payer, year, agent. Incr. = incremental cost (marginal effect: predicted mean cost with event minus predicted mean cost without). All p<0.001 except Stroke/TIA Adj. CR p=0.011 and MI Unadj. CR p=0.003. Full adjusted model in Supplemental Table 14.*

### Supplemental Table 10. Incremental 90-Day Cost of Adverse Events in the AF+DOAC Subgroup (N = 1,545 with Cost Data)

| Adverse event | N With | N Without | Mean Cost With | Mean Cost Without | Incremental Cost |
|---|---|---|---|---|---|
| Stroke/TIA | 31 | 1,514 | $20,114 | $10,416 | +$9,698 |
| Bleeding | 371 | 1,174 | $13,418 | $9,724 | +$3,694 |
| VTE (DVT/PE) | 46 | 1,499 | $19,028 | $10,352 | +$8,676 |
| Any adverse event | 429 | 1,116 | $13,710 | $9,419 | +$4,292 |

*All costs in US dollars (proxy cost). AF = atrial fibrillation; DOAC = direct oral anticoagulant; VTE = venous thromboembolism; DVT = deep vein thrombosis; PE = pulmonary embolism; TIA = transient ischemic attack.*

### Supplemental Table 11. 90-Day Peri-Procedural Cost by Anticoagulant Agent

| Agent | N | Mean Cost | Median Cost | IQR | Bleeding (%) | Stroke/TIA (%) |
|---|---|---|---|---|---|---|
| Apixaban | 973 | $10,926 | $7,624 | $4,024–$13,213 | 24.6 | 2.4 |
| Rivaroxaban | 803 | $10,059 | $6,955 | $3,640–$11,935 | 21.8 | 1.5 |
| Warfarin | 118 | $8,509 | $6,084 | $3,315–$11,781 | 28.0 | 1.7 |
| Dabigatran | 36 | $10,338 | $6,551 | $3,177–$10,580 | 22.2 | 0.0 |

*All costs in US dollars (proxy cost). IQR = interquartile range. Edoxaban (n=3) excluded due to insufficient sample size. Bleeding and stroke/TIA rates are 90-day rates using the strict event definition.*

**Supplemental Table 12. Exploratory Analysis: Association Between Peri-Procedural Bleeding and Subsequent Stroke/TIA**

**Panel A:** 90-Day Stroke/TIA Rate by Prior Bleeding Status

| Bleed Window | N Bled | Stroke/TIA in Bleeders | Stroke/TIA in Non-Bleeders | OR (95% CI) | P value |
|---|---|---|---|---|---|
| Within 7 days | 448 | 12/448 (2.7%) | 31/2,038 (1.5%) | 1.78 (0.91–3.50) | 0.09 |
| Within 14 days | 486 | 13/486 (2.7%) | 30/2,000 (1.5%) | 1.80 (0.93–3.49) | 0.08 |
| Within 30 days | 510 | 14/510 (2.7%) | 29/1,976 (1.5%) | 1.90 (0.99–3.61) | 0.05 |

**Panel B:** Temporal Relationship and Cost

| Metric | Value |
|---|---|
| Patients with stroke/TIA after a bleeding event | 13 |
| Median days from bleed to stroke/TIA | 6 |
| Mean days from bleed to stroke/TIA | 22.4 |
| Mean 90-day cost: bleed + stroke/TIA | $23,716 |
| Mean 90-day cost: bleed only | $12,065 |
| Incremental cost of combined events | +$11,651 |

*Odds ratios, 95% confidence intervals, and P values from logistic regression. Stroke/TIA ascertained using the strict event definition within 90 days of the index procedure. OR = odds ratio; CI = confidence interval; TIA = transient ischemic attack.*

**Panel C:** Multivariable Logistic Regression for 90-Day Stroke/TIA (N=2,486; 43 events)

| Variable | OR | 95% CI | P value |
|---|---|---|---|
| Age (per year) | 0.964 | 0.933–0.995 | 0.021 |
| Female sex | 0.550 | 0.274–1.104 | 0.093 |
| CHA2DS2-VASc (per point) | 1.582 | 1.239–2.019 | <0.001 |
| HAS-BLED (per point) | 1.350 | 1.009–1.806 | 0.044 |
| Procedure year | 1.115 | 0.968–1.285 | 0.131 |
| Rivaroxaban (vs apixaban) | 0.951 | 0.476–1.899 | 0.887 |
| Warfarin (vs apixaban) | 1.170 | 0.321–4.265 | 0.812 |
| Bleeding within 30 days | 1.189 | 0.595–2.375 | 0.625 |

*Logistic regression with binomial family. Dabigatran excluded due to zero stroke/TIA events (complete separation). The association between peri-procedural bleeding and subsequent stroke/TIA was not statistically significant after adjustment for baseline covariates.*

**Supplemental Table 13. Outpatient/Elective Subgroup: Peri-Procedural Event Rates and Costs (Anticoagulated Patients, N = 504)**

**Panel A:** Event Rates (Strict Definition)

| Event | 7 Days | 30 Days | 90 Days |
|---|---|---|---|
| Bleeding | 34 (6.7) | 43 (8.5) | 54 (10.7) |
| Any thromboembolic | 1 (0.2) | 8 (1.6) | 20 (4.0) |
| Stroke/TIA | 0 (0.0) | 1 (0.2) | 4 (0.8) |

**Panel B:** Incremental 90-Day Cost by Adverse event

| Adverse event | N | With Event | Without | Incremental |
|---|---|---|---|---|
| Bleeding | 37 | $11,159 | $6,957 | +$4,202 |
| *Stroke/TIA | 4 | $9,894 | $7,405 | +$2,489 |
| *VTE (DVT/PE) | 6 | $17,052 | $7,255 | +$9,798 |
| Any adverse event | 47 | $11,955 | $6,671 | +$5,284 |

*Values are n (%) for event rates and US dollars for costs. Restricted to anticoagulated patients with outpatient, ASC, or office procedure setting. * = fewer than 10 events; interpret with caution. VTE = venous thromboembolism; TIA = transient ischemic attack.*

### Supplemental Table 14. Multivariable GLM Coefficients for 90-Day Total Cost

| Variable | Coefficient | Cost Ratio | 95% CI | P value |
|---|---|---|---|---|
| Age (per year) | -0.006 | 0.994 | 0.989–1.000 | 0.034 |
| Female sex | -0.062 | 0.939 | 0.847–1.042 | 0.238 |
| CHA2DS2-VASc (per point) | 0.022 | 1.022 | 0.983–1.062 | 0.274 |
| HAS-BLED (per point) | 0.095 | 1.1 | 1.051–1.151 | <0.001 |
| ERCP (vs EMR) | 0.292 | 1.339 | 1.197–1.496 | <0.001 |
| Variceal (vs EMR) | 0.157 | 1.17 | 1.013–1.350 | 0.032 |
| Drainage (vs EMR) | 0.509 | 1.663 | 1.305–2.120 | <0.001 |
| POEM (vs EMR) | -0.243 | 0.784 | 0.499–1.234 | 0.294 |
| Medicare (vs Commercial) | -0.118 | 0.889 | 0.786–1.005 | 0.06 |
| Medicaid (vs Commercial) | -0.251 | 0.778 | 0.689–0.878 | <0.001 |
| Procedure year | 0.017 | 1.017 | 0.997–1.037 | 0.096 |
| Rivaroxaban (vs Apixaban) | -0.04 | 0.961 | 0.868–1.065 | 0.449 |
| Warfarin (vs Apixaban) | -0.272 | 0.762 | 0.619–0.937 | 0.01 |
| Dabigatran (vs Apixaban) | -0.011 | 0.99 | 0.696–1.407 | 0.954 |

*Generalized linear model with gamma family and log link. N=1,925. Cost ratio = exp(coefficient); values >1.0 indicate higher costs. Reference categories: EMR for procedure, Commercial for payer, Apixaban for anticoagulant agent.*

**Supplemental Figure 1.** Patient study flow diagram

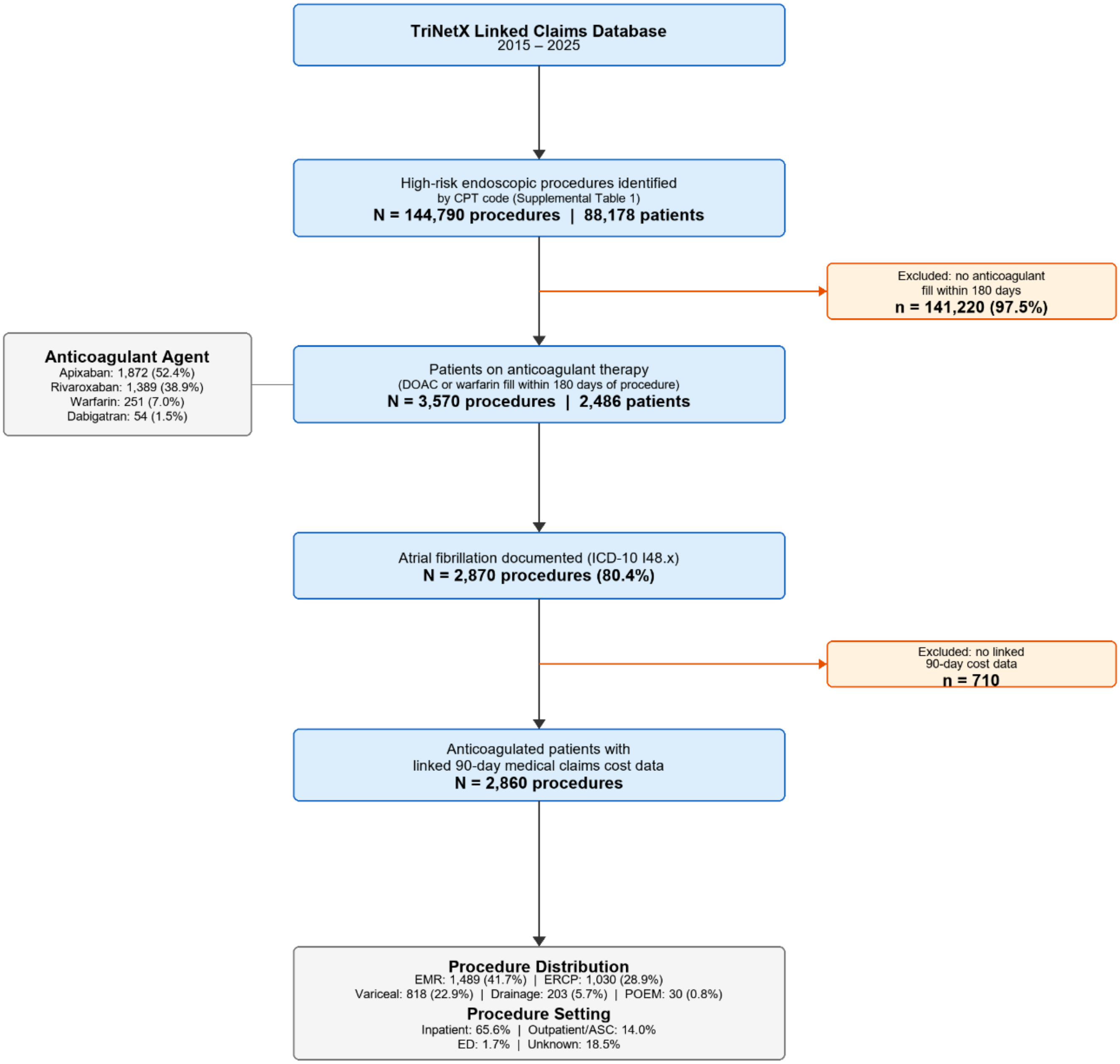

**Supplemental Figure 2.** Peri-procedural event rates in anticoagulated patients by time window (7, 30, and 90 days) using the strict event definition.

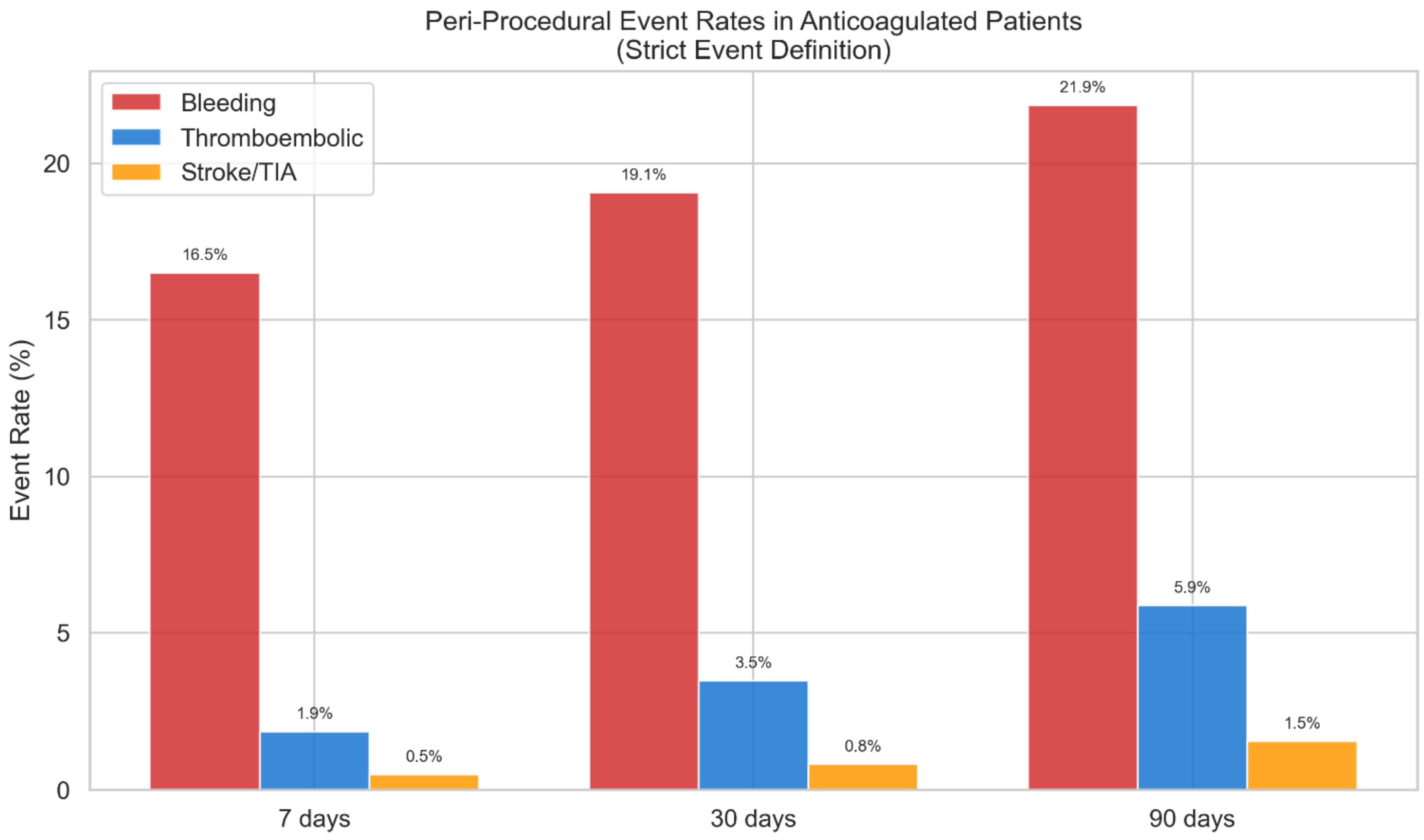
